# How many infection control staff are needed in acute care hospitals? A Delphi approach

**DOI:** 10.64898/2025.12.09.25340557

**Authors:** Marlies Mulder, Maarten Sarink, Gerrie Stoffer, Miriam Mes, Els van Oorschot, Laura van Dommelen, Andreas Voss, Juliëtte A. Severin, Karin-Ellen Veldkamp, Rosa van Mansfeld

## Abstract

**Introduction:** The Dutch recommendation on infection prevention and control (IPC) staffing in acute care hospitals from 2007 is outdated due to evolving hospital care, including shorter admissions, more complex and day-care procedures, more vulnerable patients, increasing antimicrobial resistance, and enhanced regulatory demands. Therefore, an updated staffing norm for IPC is needed.

**Methods:** Minimum weekly hours required for IPC activities was determined by Delphi method across three model hospitals: academic, large non-academic, and small non-academic. Four questionnaire rounds were conducted among IPC practitioners (IPCP) and clinical microbiologists (CM). Staffing needs per role and hospital type were calculated. After three rounds a core expert team focus group formulated a new full time equivalent (FTE) norm which was proposed in the final round.

**Results:** For academic hospitals, 100% consensus was achieved for a minimum of 0.15 FTE CM and 1.23 FTE IPCP per 5000 annual hospital admissions, plus 0.05 FTE CM and 0.41 FTE IPCP per 5000 annual day admissions, respectively. For non-academic hospitals, 92% supported the proposed norm for CM (same values), and 89% agreed with the proposed norm for IPCP: 1.10 FTE per 5000 hospital admissions and 0.37 FTE per 5000 day admissions.

**Conclusion:** A new consensus-based staffing norm, endorsed by most Dutch IPC professionals, recommends an increase in IPCP. This reflects increased demands on IPC teams and suggests diversification of professionals working in IPC teams, not accounted for in the previous norm. This minimum norm is needed to effectively protect patients and healthcare workers from infections.

## Introduction

The COVID-19 pandemic and the continued rise in antimicrobial resistance have brought renewed and widespread attention to the importance of infection prevention and control (IPC) [1]. Despite this, the subject of IPC staffing remains relatively underexplored, with most research originating from the United States [2-4]. One of the first studies on this topic, the SENIC study (1985), recommended one IPC practitioner (IPCP) per 250 hospital beds [3]. More recent studies have reported staffing ratios ranging from one IPCP per 152 beds to one per 69 beds [4]. However, IPC staffing requirements depend heavily on defined responsibilities, healthcare setting (e.g., acute vs. long-term care), and organizational context, which vary internationally.

In the Netherlands, the Quality Guideline for Infection Control in Hospitals (KRIZ), revised in 2021, offers clear recommendations regarding the organization and responsibilities of IPC teams in acute care settings [5]. The Dutch guideline on IPCP staff, however, dates back to 2007. It recognized that bed numbers inadequately reflect IPC workload in Dutch hospitals due to high patient turnover, proposing instead the number of admissions as a more accurate metric. That norm recommended one full-time equivalent (FTE) IPCP per 5,065 admissions and one FTE clinical microbiologist (CM) per 22,941 admissions—a benchmark that has since been used by the Health and Youth Care Inspectorate. [6].

Over the past two decades IPC teams in acute care hospitals have faced growing demands driven by increasing care complexity and medical innovations. Also, more stringent regulatory and accreditation requirements, next to more patient and public awareness on patient safety, increase the workload of IPC teams. New epidemiological insights, as such gained by whole-genome sequencing, the possibilities associated with electronic patient files, and “big data” have changed the demands on skillsets needed within IPC teams. More recently, sustainability and how to find an optimum between safety and environmental costs have had a growing impact on infection control workload [7]. Finally, the organization of acute care has evolved towards shorter stays and increased numbers of day admissions, further intensifying workload.

Taken together, these developments have rendered the 2007 staffing norm outdated. The aim of this study was to establish consensus on a revised IPC staffing norm that reflects current needs, through a structured Delphi process and expert team focus group discussions. This updated norm is intended to guide policy-makers and hospital IPC leadership in ensuring adequate and sustainable IPCP staffing.

## Methods

Between July 2022 and July 2024, we conducted a Delphi study; an iterative, multistage process designed to systematically gather and refine expert opinions through multiple rounds of anonymous feedback, with the goal of achieving consensus [8]. The study was conducted in accordance with the CREDES (Conducting and Reporting of Delphi Studies) guidelines for methodological rigor and transparency [9]. It was initiated by the Dutch Society of Medical Microbiology (NVMM) and the Dutch Society for Hygiene and Infection Prevention in Healthcare (VHIG). A detailed overview of the study procedure is provided below and illustrated in Figure 1. A waiver for ethic approval was obtained from the Institutional Review Board at Amsterdam UMC (2022.0455).

**Figure 1.**
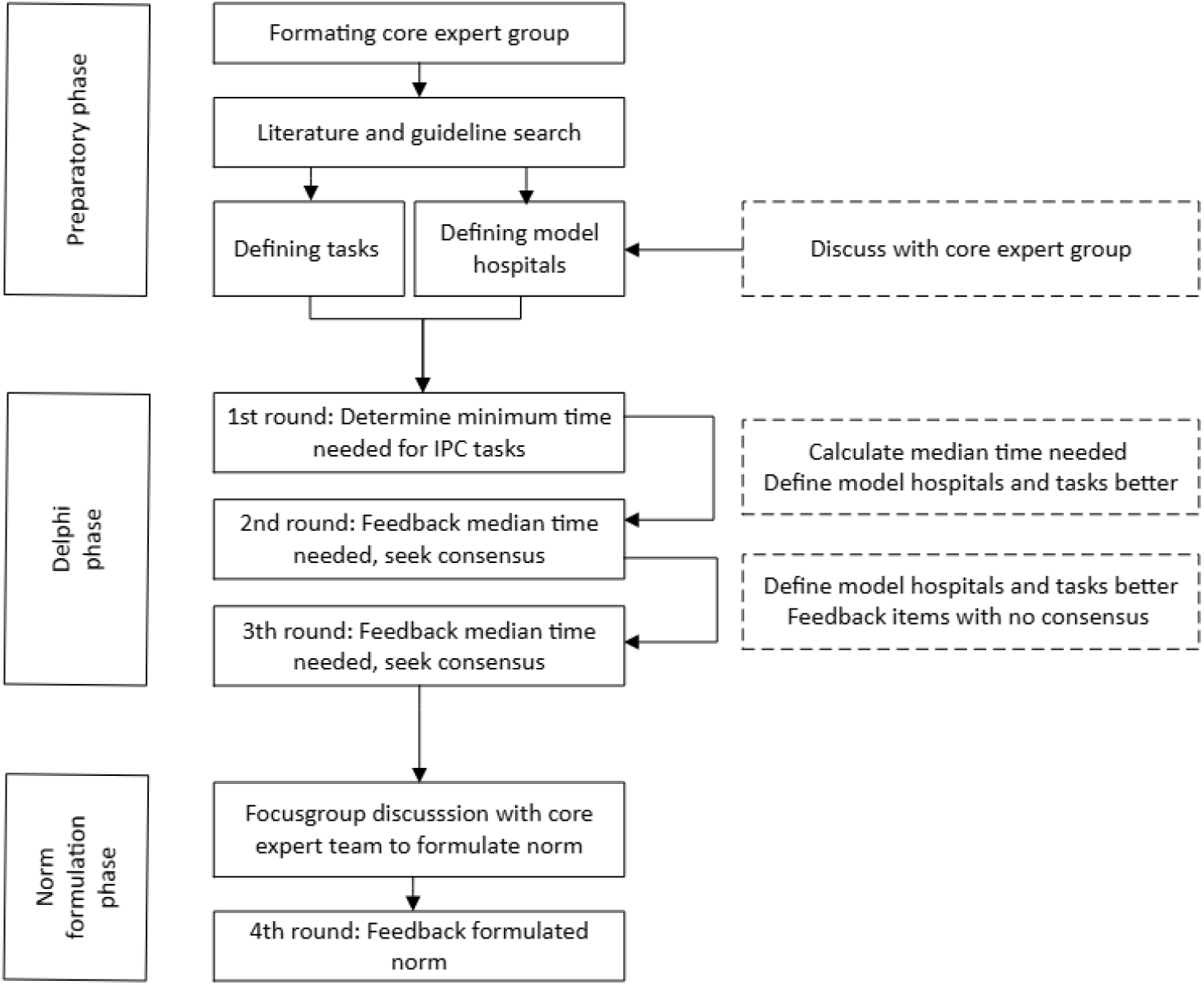
Flowchart of process stages

### Study design

First, three standard model hospitals were defined, and a comprehensive list of IPC tasks was compiled (described in detail below). These preliminary definitions were reviewed and refined during a focus group discussion with the core expert team (further described below). Subsequently, a series of Delphi rounds was conducted to search for consensus.

The first round was used to collect input on both the current and the minimum required staffing (in hours per week) needed to perform specific IPC tasks. In the next rounds, participants were presented with the median values from round one and were asked to indicate their level of agreement on a 5-point Likert scale and to provide justification or comments. After each round, the results were evaluated by the core expert team, who discussed potential modifications to the task descriptions or model hospital definitions for the next round. When no further changes in consensus were expected, the core expert team defined a proposed norm based on the aggregated findings from the previous rounds. A final round was aimed to establish consensus on this proposed norm.

### Core expert team en expert panel selection

A core expert team was formed by inviting leading professionals in the field, through both professional associations (VHIG and NVMM) to contribute to the design and objectives of the study and to participate in online focus group discussions. This team consisted of four CM and three IPCP, representing three academic and three non-academic Dutch hospitals.

The broader expert panel for the Delphi study comprised members of the NVMM and VHIG. Potential participants were contacted via email through their respective associations and asked to designate one CM and one IPCP per IPC department in each Dutch hospital.

### Infection control tasks

A comprehensive list of tasks covering the tasks of IPC teams was developed based on the 2007 infection prevention norm and the updated KRIZ guideline [5, 6]. The final set of tasks was defined and validated by the core expert team, resulting in 13 key task areas: surveillance; infection prevention policy; outbreak management and preparedness; advisory and consultative services; education and training; auditing and monitoring; quality management and policy development; in-service training; availability outside regular working hours; participation in committees; research; external consulting; and miscellaneous activities (Supplementary Table 1).

### Model hospitals

To ensure the applicability of the proposed norm and enable fair comparison across participants, respondents were asked to project their local IPC workload onto a hypothetical, standardized model hospital. Three model hospitals were introduced, reflecting typical hospital types in the Netherlands: small regional hospitals, large regional hospitals, and academic tertiary care centres. To define the characteristics of these model hospitals, a random selection of 15 to 20 small and large regional hospitals was made, along with all seven academic medical centres. Publicly available data were collected on key hospital metrics, including annual inpatient admissions, outpatient visits, number of operating theatres, staff size, and level of care. Based on these data, average values were calculated to establish representative profiles for each model hospital (Supplementary Table 2).

### Questionnaires

Participants of the expert panel were invited to complete an online questionnaire using Survalyzer, a platform for designing and analysing surveys. CM and IPCP completed the survey either separately or jointly. Separate responses were used within their own groups; joint responses applied to both. Both could respond on behalf of other IP staff. The survey link was distributed via email by the VHIG and NVMM to all hospital IPC departments in the Netherlands.

### First round

Participants were asked to estimate the minimum time (in hours per week) required for CM, IPCP and other IPC team staff to perform the predefined tasks within a selected model hospital based on the current situation of their hospital. ‘Other IPC staff’ includes for instance epidemiologists, data analysts, assistants and quality officers. Staff in training, sterile medical devices specialists and endoscope reprocessing specialists were explicitly excluded. Where relevant, time estimates from the 2007 recommendation were provided for comparison. The same questions were asked for all staff roles. Medians and interquartile ranges (IQR) were calculated per task, per role, and per hospital type.

### Second round and third round

All participants of the first round were asked to assess their agreement with the median time estimates for each task and role using a five-point Likert scale (1 = strongly disagree, 5 = strongly agree). If they disagreed, they were asked to provide justification.

For tasks without consensus, descriptions were revised based on round-two feedback in the third round. Participants were again asked to indicate their agreement using the same scale. Consensus in these rounds was defined as ≥75% of participants scoring 3 (neutral) or higher.

### Norm formulation by the core expert team

Results from rounds two and three were reviewed by the core expert team, who refined the estimates and translated them into a generalizable norm. Several calculation models were considered (e.g., per number of admissions or per number of staff), and consensus was reached on the most suitable determinants through a panel discussion.

### Fourth round

The proposed norm was submitted for approval to all Dutch CM and IPCP via their professional associations. Each hospital could cast one vote per CM and IPCP. When the survey was completed together by a CM and an IPCP, the response was counted as one vote for each group. Consensus was defined as ≥75% approval.

## Results

### Demographics of expert panel

The initial questionnaire was completed by 68 respondents from 58 different hospital locations. The Netherlands has 68 hospital organizations, including 7 university medical centers, spread across 113 locations. Some locations have separate IPC teams, which could submit individual responses. Most questionnaires in the first round were completed by IPCPs (n = 40) (Table 1). Although the second and third round questionnaires were sent to all initial participants, response rates declined in subsequent rounds. Additionally, an increasing number of CM and IPCP respondents completed the survey together. Following the formulation of the new staffing norm by the core expert team, the proposed norm was distributed to all CM and IPCP across Dutch hospitals. This final questionnaire was completed by 96 respondents, representing 70 hospital locations from 55 hospital organizations. Of these, 26 were completed by CMs, 38 by IPCPs (including team leads with an IPC background), and 32 together by a CM and IPCP (Table 1).

**Table 1:**
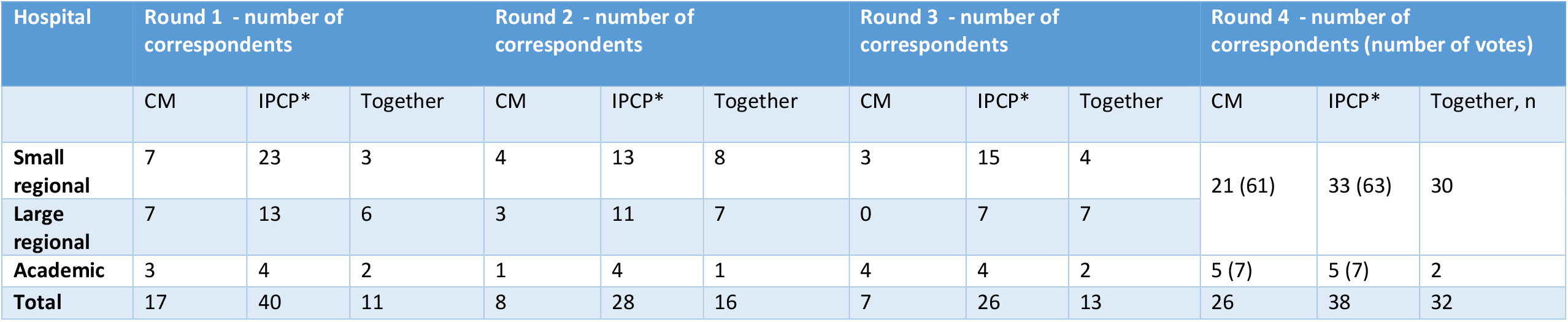
Number of respondents per hospital type. Table 1 shows the total of completed questionnaires per hospital type (small regional, large regional and academic and per group (clinical microbiologist (CM), infection control practitioner (*IPCP; includes infection control heads, managers with IPC background) and CM and IPCP together). For round 1, all CM specialized in infection control and all IPCP were invited. For round 2 and 3, all participants that responded in round 1 were invited. For round 4, again, all CM specialized in infection control and all IPCP were invited (in between brackets the number of votes for this group: e.g. for CM of small regional hospitals 21 from CM + 30 from CM and IPCP who filled out the questionnaire together = 61 votes).

### First round

The definitions of the model hospitals and list of IPC tasks (Supplementary Tables 1 and 2) were established through panel discussions within the core expert team.

The main outcome of the first Delphi round was the median time needed per IPC task item per model hospital per IPC professional group. Remarkably, most respondents estimated to need more time to perform IPC activities on the list of IPC tasks than was currently available in their institutions. Additionally, many participants indicated that they required more detailed information about the model hospitals to reliably translate their local context to the hypothetical setting. Several respondents also reported difficulties in determining which specific responsibilities fell under each IPC task item. The task “availability outside office hours” was excluded from further analysis, as it was found to vary widely between institutions and did not yield generalizable or useful input.

Consequently, this task is not reflected in the final staffing norm.

### Second round

More detailed information was given on the characteristics of the model hospitals (Supplementary Table 2) as well as on the scope and content of the infection prevention tasks, specifically that time for communication (website, patient information letters, meetings, making posters etc.) is incorporated in several tasks.

For the small and large regional model hospitals, the hours reported for “assistant” and “other staff” were combined with those of the IPCP, as the individual numbers were too low to analyse separately. For the academic model hospital, three staff categories were retained: CM, IPCP, and “other staff” (including assistant IPCs and other IPC team members).

Consensus was achieved for 16 out of 77 tasks (21%). For the remaining tasks, there was no clear agreement—responses varied, with some participants indicating a need for more time, while others estimated less time would be sufficient.

> *“Very dependent on the number of outbreaks. The Infection Prevention Department handles all preparations and sometimes also chairs the Outbreak Management Teams (OMTs)*.*”* [disagree time needed for outbreak management and preparedness, IPCP large regional hospital]
>
> *“Setting up and maintaining surveillance takes time. By keeping it in your own hands, you can respond well to local issues. This requires a certain basic investment of time, which can increase if problems arise. For that reason, the difference in hours between smaller regional hospitals on the one hand and academic and large regional hospitals on the other hand cannot be that large in practice*.*”* [disagree time needed for surveillance AM & IPCP from small regional hospital]
>
> *“In an outbreak situation, a significant number of hours are required for this, but outside of outbreaks, the minimum requirements are too high*.*”* [disagree outbreak management and preparedness, AM from large regional hospital]

Some respondents noted additional or specialized tasks specific to their local context, such as managing high-level isolation units, overseeing large construction projects, participating in regional initiatives, or supporting affiliated long-term care facilities.

### Third round

In the third round it was emphasized that the objective was to establish a norm reflecting the absolute minimum of hours per week required to perform a set of basic IPC tasks in acute care hospitals. Further clarification was provided regarding context and task descriptions: The hospital serves an average Dutch population, has a digital surveillance system, participates in national HAI registration (at least one module), is in good structural condition and has one infection prevention specialist in training.

Consensus was reached on 56 out of 77 tasks (73%) (Supplementary Table 3). However, there was no consensus on the total minimum weekly hours required for IPC staff across all roles—except for CM in the academic hospital model. Most comments came from panel members who felt the proposed median time was insufficient.

> *“Developing policies is the same [workload] for every hospital, whether it is large or small. So the absolute minimum should be the same for every type of hospital. Academic hospitals may need more, because they have more specific departments. But [time needed in] small regional hospitals should be equal to large regional hospitals”* [disagree on time for on infection prevention policy, IPCP from small regional hospital]
>
> *“It’s not just about guidelines, but also about fine-tuning policy, adapting it to the current standard, consulting with stakeholders to increase support*.*”* [disagree on time for on infection prevention policy AM& IPCP from large regional hospital]
>
> *“In the coming years, guidelines are expected to change more often and implementing hospital-wide policy now requires more effort*.*”* [disagree on time for infection prevention policy, AM and IPCP from small regional hospital]

### Formation of the norm

Following two focus group discussions, the core expert team agreed to propose a new minimum norm for basic infection control activities, based on the number of hospital admissions and day admissions, using a 3:1 weighting ratio (hospital admissions vs. day admissions). Given the minimal differences between small and large regional hospitals, their norms were merged into a single norm. To further streamline the model, a single norm was defined for CM and one for all other IPC team members, separately for academic and non-academic hospitals.

During the expert panel discussion—focused on total time requirements rather than individual tasks—minor adjustments were made to the estimated minimum time needed for IPC staff, incorporating participant feedback. These adjusted estimates formed the basis for the proposed norm, calculated using the characteristics of the model hospitals.

This proposed norm for CM for all hospitals was: 0.15 FTE CM per 5,000 hospital admissions per year, plus 0.05 FTE CM per 5,000 day admissions per year. For IPCP and other IPC team members in academic hospitals: 1.23 FTE IPCP per 5,000 hospital admissions per year, plus 0.41 FTE IPCP per 5,000 day admissions. For non-academic hospitals: 1.10 FTE IPCP per 5,000 hospital admissions per year, plus 0.37 FTE IPCP per 5,000 day admissions. (Table 2)

**Table 2:** Proposed norm for number of FTE for CM and IPCP per number of hospital admissions and day admissions.

|  | CM | IPCP |  |
| --- | --- | --- | --- |
|  |  | Non-academic | Academic |
| Per 5000 hospital admissions | 0.15 | 1.10 | 1.23 |
| Per 5000 day admissions | 0.05 | 0.37 | 0.41 |

FTE = fulltime-equivalent , CM = clinical microbiologist, IPCP = infection prevention and control practitioner

### Fourth round

In the final round, the process and key considerations of the expert panel were explained, and the proposed minimum staffing norm was presented for approval. Respondents from academic hospitals showed full agreement (100%) with the proposed norm. For non-academic hospitals, 92% agreed with the norm for CM, and 89% agreed with the norm for IPCP and other IPC staff.

## Discussion

As hospital care evolves, the demands on IPC teams have steadily increased, necessitating a revision of existing staffing norms. The previous Dutch guideline from 2007 is outdated. Between 2022 and 2024 we used a Delphi method to reach expert consensus on the minimum time required by IPCP and CM for core infection control tasks. The final proposed norm received broad support from IPC professionals across Dutch acute care hospitals.

While the recommended minimum FTE for CM remained comparable to the 2007 standard, the new norm indicates a 50–60% increase in FTE for IPCP. This rise reflects both the increased needs due to the growing scope and complexity of IPC work and the inclusion of additional team roles, such as epidemiologists, team managers, data managers, and secretarial staff, who were not considered in the previous norm [10, 11]. Robinson et al investigated what an optimal IPC service needs and found that adequate staff resources and a trained competent IPC team are major components for this [11]. A recent U.S. position paper similarly calls for raising standards for IPC programs and proposes staffing calculators (APIC Staffing Calculator) and frameworks to assess institutional needs [12], which unfortunately is only available for members.

Importantly, our new norm covers only basic infection prevention activities. In the second and third Delphi rounds, responses varied considerably. Much of this variation was caused by differences in hospital structure, local task assignment, and the challenge of extrapolating local circumstances to the model hospitals. Some hospitals reported performing tasks beyond the basic IPC scope—such as vaccinations, needlestick injury follow-up, and MRSA screening for staff—or providing IPC services to affiliated long-term care facilities. Others had highly specialized functions, such as managing viral hemorrhagic fever units or had to deal with large newbuilt projects. This is explicitely not included in this norm and would need extra formation. Additionally, IPC workload is subject to seasonal or episodic surges, such as during outbreaks or pandemics which also require more formation. Very small hospitals also noted that some baseline investment—like policy development and education— is always needed, regardless of hospital size, making the relative burden higher, and they will likely need more time than the proposed norm. There was also notable variation in IPC team composition. In some hospitals, tasks are distributed among dedicated support staff (e.g., data managers or infection control epidemiologists), while in others, IPC practitioners are responsible for all duties.

Although these differences were addressed by including all team members in the IPCP norm, this highlights the importance of flexible, competency-based team composition.

The Delphi method provides a structured technique to reach expert consensus on complex topics [13]. While it enables broad expert input, it has limitations. Selection bias and differential willingness to participate may influence outcomes. However, IPC teams from over 80% of Dutch acute care hospitals participated in both the first and final rounds, enhancing the representativeness of our findings. Some risk of conformity bias exists in multi-round surveys, but anonymity and opportunities for free-text feedback mitigated this. Dropout across rounds did occur, though updates via national meetings helped maintain engagement and IPC teams from all Dutch hospitals were again invited to participate in the final round. Finally, this Delphi method offers expert-informed estimates for the Dutch situation. The results might not be easily extrapolated to other type of care facilities or other countries with different organization of care and responsibilities. However, the methodology is transferable and adaptable to other countries and healthcare settings. This enhances the global relevance of our findings beyond the Dutch context.

## Conclusion

The role of IPC teams in acute care hospitals has evolved significantly, requiring greater flexibility, multidisciplinary collaboration, and stronger workforce capacity. This study presents a consensus-based minimum staffing norm for IPC teams in Dutch acute care hospitals, based on hospital admissions and day admissions. The norm reflects the increasing complexity and scope of IPC activities and was widely supported by IPCP and CM. This new norm can serve as a reference for hospital management, IPC teams, and national regulatory authorities such as the Health and Youth Care Inspectorate. Amidst increasing demands and workforce shortages, this work supports more resilient and effective IPC services in the future.

## Data Availability

All data produced in the present study are available upon reasonable request to the authors

## Notes

### Competing Interest Statement

Juliette Severin:
Board member of Dutch Society for Medical Microbiology (Unpaid) Chair of IPC working group of ISAC (Unpaid) Member of Regional Coordination Team of the IP & AMR Care Network Southwestern Netherlands (Payment to Erasmus MC) Andreas Voss Chair of Dutch IPC guideline group (paid) Steering Committee of EUCIC (unpaid) Other authors: none

### Funding Statement

This study did not receive any funding

### Summary of Updates

Title from: How many infection control staff is needed in acute care hospitals? A Delphi approach. To How many infection control staff are needed in acute care hospitals? A Delphi approach.

