## Supplementary materials for "How many infection control staff are needed in acute care hospitals? A Delphi approach"

**Supplementary Table 1: Infection control tasks and the further clarification that was given in round 2.**

| Task | Further clarification round 2 |
|---|---|
| **Surveillance** | **Surveillance is defined as:**<br>a. Detection, registration, and analysis of healthcare-associated infections, multidrug-resistant organisms (MDROs), and other relevant microorganisms.<br>b. Discussion of results with stakeholders and responsible parties, providing recommendations for improvement measures, and monitoring follow-up actions.<br>c. Management and maintenance of ICT systems that collect surveillance data. |
| **Infection prevention policy** | **Infection prevention policy is defined as:**<br>The development, maintenance, implementation, and evaluation of infection prevention guidelines for the organization, covering areas such as precautionary measures, isolation, infectious diseases, operating rooms, and surveillance. This also includes the creation of new guidelines and any policy adjustments resulting from Green Teams. |
| **Outbreak management and preparedness** | **Outbreak management and preparedness is defined as:**<br>a. Identifying (potential) outbreak situations or other infection prevention incidents and taking on a coordinating role in response.<br>b. Developing protocols to enable timely recognition of and response to outbreaks.<br>c. Conducting emergency preparedness and outbreak response training. |
| **Advisory and consultative services** | **Advisory and consultation tasks include:**<br>a. Providing solicited and unsolicited advice, responding to questions regarding infection prevention policy, and documenting the advice given.<br>b. Matters related to cleaning, disinfection, and sterilization. This includes, among other things, the development of disinfection policies and endoscope management plans, information exchange with departments such as departments for sterilization of medical instruments, and the evaluation and follow-up of related policies.<br>c. Advising on new construction and renovation projects. |

| | |
|---|---|
| | d. Advising on procurement and implementation processes (e.g., the acquisition and use of equipment and materials).<br>e. Advising on water and air quality management. |
| **Education and training** | **Education and training include:**<br>a. Developing training programs (e.g., e-learning).<br>b. Providing training and education to healthcare workers.<br>c. Supervising residents and infection prevention trainees.<br>d. Teaching students. |
| **Auditing and monitoring** | **Auditing and monitoring include:**<br>a. Developing audit plans, questionnaires, and checklists.<br>b. Conducting QuickScans and audits, and preparing reports.<br>c. Performing follow-up activities, such as monitoring improvement plans and compliance. |
| **Quality management and policy development** | **Quality management system and policy cycle include:**<br>a. Developing, maintaining, evaluating, and improving the quality management system (PDCA cycle).<br>b. Responding to incident reports (VIM), and formulating improvement actions.<br>c. Preparing multi-year policies, annual plans, annual reports, and periodic progress reports. |
| **In-service training** | **In-service training:**<br>Attending symposia and conferences, as well as participating in courses to keep knowledge up-to-date and to stay informed about new guidelines and innovations in infection prevention. |
| **Availability outside regular working hours** | **Availability for questions and advice on infection prevention policy to staff outside office hours.**<br>The actual number of hours that the clinical microbiologist (CM) and infection prevention and control practitioner (IPCP) are required to actively work outside office hours. |
| **Participation in committees** | **Participation in committees include:**<br>Participation in and responsibilities for internal and external committees, such as the infection prevention committee, |

| | |
|---|---|
| | quality committee, regional infection prevention working groups, and national committees involved in guideline development. |
| **Scientific research** | **Scientific research includes:**<br>Conducting relevant scientific research or contributing to such research within the scope of infection prevention. |
| **External counselling** | Not further specified |
| **Miscellaneous activities** | Not further specified; tasks mentioned in the comments were included under one of the categories above. |

**Supplementary Table 2:** **Characteristics of the three different model hospitals to which participants needed to extrapolate their data.**

| | **Model academic hospital** | **Model large regional hospital** | **Model small regional hospital** |
|---|---|---|---|
| **Total number of admissions per year** | 50000 | 50000 | 25000 |
| **Clinical admissions** | 25000 | 25000 | 12500 |
| **Day admissions** | 25000 | 25000 | 12500 |
| **Number of outpatient visits** | 80000 | 130000 | 75000 |
| **Operating rooms** | 25 | 15 | 8 |
| **Number of employees** | 11000 | 4000 | 2300 |

**Supplementary Table 3: Minimum number of hours per week for the different professional groups after round 3**

| Minimum number of hours per week. Median (IQR) | Small regional hospital | | Large regional hospital | | Academic hospital | | |
|---|---|---|---|---|---|---|---|
| | CM [IQR] | IPCP and other [IQR] | CM [IQR] | IPCP and other [IQR] | CM [IQR] | IPCP [IQR] | other [IQR] |
| **Surveillance** | 1.5 [1.0-2.0] | 16.0 [12.0-32.0] | 3.0 [2.0-5.0] | **41.0 [32.1-54.5]** | **3.0 [1.3-3.0]** | 24.0 [10.0-36.0] | 14.0 [4.0-24.0] |
| **Infection control policy** | 2.0 [1.1-3.0] | 16.0 [10.0-20.0] | 3.0 [2.8-6.5] | 26.0 [20.0-36.0] | 5.0 [4.0-12.5] | 29.0 [20.0-36.0] | 4.0 [0.0-13.8] |
| **Outbreak management en preparedness** | **2.0 [1.0-2.6]** | **7.0 [4.5-14.5]** | 4.0 [3.0-6.5] | 12.3 [7.5-22.3] | **3.5 [2.0-4.0]** | 8.5 [8.0-18.4] | 4.0 [0.0-6.0] |
| **Advisory and consultative services** | 2.0 [1.0-3.3] | 28.0 [20.0-50.0] | 4.0 [2.0-7.0] | **56.0 [40.3-61.5]** | **8.0 [3.0-8.0]** | **60.0 [40.0-72.0]** | 21.0 [0.0-40.0] |
| **Education and training** | **0.7 [0.5-1.0]** | **11.0 [5.0-14.5]** | 1.0 [0.5-3.0] | 20.0 [10.3-29.0] | **2.0 [1.0-3.8]** | 36.0 [12.0-50.0] | 3.5 [0.0-10.0] |
| **Auditing and monitoring** | **0.9 [0.5-1.0]** | 10.0 [8.0-15.5] | 1.2 [0.4-2.0] | **17.5 [14.3-20.4]** | **1.0 [0.5-1.0]** | 10.0 [8.0-23.1] | 1.0 [0.0-3.8] |
| **Quality management and policy development** | 1.0 [0.5-2.0] | 6.0 [4.0-10.0] | 2.0 [1.0-3.3] | 12.5 [6.5-18.8] | **3.0 [2.0-4.0]** | 8.0 [5.0-17.2] | 2.0 [2.0-4.0] |
| **In-service training** | **1.0 [0.5-1.9]** | 6.0 [4.0-8.0] | 3.0 [0.8-4.0] | **12.5 [7.0-16.8]** | **2.0 [1.3-2.0]** | **10.0 [8.9-12.0]** | **2.0 [0.0-3.1]** |
| **Participation in committees** | **1.8 [1.0-2.7]** | 8.0 [5.0-11.0] | 4.0 [2.0-6.0] | **11.8 [4.3-15.8]** | **4.0 [3.0-6.0]** | **8.9 [8.0-14.0]** | 1.0 [0.0-4.0] |
| **Scientific research** | **0.5 [0.0-1.0]** | 1.0 [0.0-2.0] | 2.0 [0.0-2.6] | 4.0 [1.3-8.3] | **6.0 [4.0-8.0]** | 2.0 [1.0-16.0] | 4.0 [0.0-23.0] |
| **TOTAL** | 14.1 [11.3-19.7] | 122 [81-175] | 34 [17.8-45.1] | 239 [190-285] | **41 [32-54]** | 216 [159-270] | 80 [33-131] |

Minimum number of hours per week per professional group (CM = clinical microbiologist, IPCP = infection prevention and control practitioner, other = all other staff in the infection and control department, such as assistants, epidemiologists, data scientists). Consensus was reached on the **bold green** numbers.
